# Quantifying superspreading for COVID-19 using Poisson mixture distributions

**DOI:** 10.1101/2020.11.27.20239657

**Authors:** Cécile Kremer, Andrea Torneri, Sien Boesmans, Hanne Meuwissen, Selina Verdonschot, Koen Vanden Driessche, Christian L. Althaus, Christel Faes, Niel Hens

**Author notes:** joint first author (Cécile Kremer, Andrea Torneri). corresponding author, Agoralaan Gebouw D, 3590 Diepenbeek, Belgium.

## Abstract

The number of secondary cases is an important parameter for the control of infectious diseases. When individual variation in disease transmission is present, like for COVID-19, the number of secondary cases is often modelled using a negative binomial distribution. However, this may not be the best distribution to describe the underlying transmission process. We propose the use of three other offspring distributions to quantify heterogeneity in transmission, and we assess the possible bias in estimates of the offspring mean and its overdispersion when the data generating distribution is different from the one used for inference. We find that overdispersion estimates may be biased when there is a substantial amount of heterogeneity, and that the use of other distributions besides the negative binomial should be considered. We revisit three previously analysed COVID-19 datasets and quantify the proportion of cases responsible for 80% of transmission, *p*_80%_, while acknowledging the variation arising from the assumed offspring distribution. We find that the number of secondary cases for these datasets is better described by a Poisson-lognormal distribution.

## Introduction

For any communicable disease, the basic reproduction number, *R*_0_, denotes the average number of secondary cases a single infected individual generates in a completely susceptible population [1, 2]. The basic reproduction number is considered to be of constant value among population members or specific population groups. However, for person-to-person transmitted infections, a complex combination of host, pathogen, and environmental factors defines the transmission potential of an infected individual, i.e. the number of other individuals a case infects during their infectious period [3, 4]. It has been shown that, for a given *R*_0_, both the probability that an epidemic will occur and the subsequent course of the epidemic are affected by individual variation in transmission [3]. Variation in disease transmission may raise the existence of ‘superspreaders’ who infect substantially more individuals than others. When superspreading plays an important role during the epidemic, a relatively small part of infected cases will be responsible for most of the transmission, while many cases do not transmit the disease at all. Furthermore, when variation in disease transmission is present, large outbreaks can occur even if *R*_0_ is less than one. To account for this heterogeneity, the individual number of secondary cases can be described by a random variable, whereas *R*_0_ represents the expected value for an entire susceptible population.

The transmission potential of infected individuals can be defined as a combination of their biological infectiousness (i.e. viral shedding) and their contact behaviour [5]. It is reasonable to assume that individuals with a higher viral shedding will be more likely to transmit the infection given a contact. In addition, for a fixed level of viral shedding, infectious individuals with a higher contact rate will be more likely to generate secondary cases. Regarding SARS-CoV-2, significant individual variation in viral shedding has been reported [6] and it has been argued that small aerosols exhaled during normal speech may serve as an important transmission route [7]. Vuorinen *et al*. [6] investigated the possibility of SARS-CoV-2 transmission by inhalation of virus-containing aerosols, by examining a high-risk scenario where an infected individual coughs within a public indoor space. They found that there was an elevated risk of infection in case of lengthy exposure in a confined space with at least one infected individual. These results are in line with those from another study which has indicated that the virus may remain infectious as an aerosol for at least three hours [8]. Of course, not only individual characteristics such as viral shedding but also environmental characteristics such as insufficient ventilation contribute to the possibility of a superspreading event (SSE) [9].

Lloyd-Smith *et al*. [3] addressed heterogeneity in transmission by using the concept of an individual reproduction number as a random variable that represents the expected number of secondary cases caused by a particular infected individual. In that framework, SSEs are important realizations from the right-hand tail of the distribution of the individual reproduction number. Most studies investigating the amount of heterogeneity in disease transmission have assumed a Poisson process with rate given by the individual reproduction numbers, assumed to follow a Gamma distribution, resulting in a negative binomial offspring distribution [3, 10]. In this way, heterogeneity has often been quantified using the *k* parameter, with *k* the negative binomial dispersion parameter. This has allowed comparison between studies, where lower values of *k* indicate increased heterogeneity in transmission, and thus possibly a larger amount of superspreading.

Based on this framework, a substantial amount of individual variation in the transmission of SARS-CoV-2 has been described, though large differences were found between different studies. Bi *et al*. [11] used a negative binomial distribution to describe superspreading in the COVID-19 outbreak in Shenzhen, China, and found that about 9% of all cases were responsible for 80% of transmission. Riou and Althaus [12] estimated the negative binomial dispersion parameter *k* to have a median of 0.54 (90% HDI 0.014 - 6.95), with simulations suggesting that very low values of overdispersion (<0.1) are less likely. Adam *et al*. [13] estimated the overall mean number of secondary cases to be 0.58 (95%CI 0.45 - 0.72) with a dispersion parameter *k* of 0.43 (95%CI 0.29 - 0.67) in Hong Kong, indicating that 19% of cases were responsible for 80% of all local transmission. Similarly, Endo *et al*. [14] have used a branching process model where the number of secondary cases was assumed to follow a negative binomial distribution. Assuming *R*_0_ to be 2.5, they estimated the dispersion parameter *k* to have a median of 0.1 (95% CrI 0.05 - 0.2), resulting in 80% of secondary cases being caused by about 10% of infectious cases and implying that large transmission events should be prevented in order to contain epidemic spread. Laxminarayan *et al*. [15] estimated the negative binomial dispersion parameter *k* to be 0.51 (95%CI: 0.49-0.52) using a large contact tracing dataset from two Indian states. Based on detailed contact tracing data from Hunan, China, Sun *et al*. [16] found that 15% of cases were responsible for 80% of transmission, and a negative binomial dispersion parameter *k* of 0.3. Lau *et al*. [17] found that superspreading was widespread across space and time, with an increasing presence towards later stages of the investigated outbreaks, highlighting the importance of maintaining social distance measures. They also found that about 2% of the most infectious cases were directly responsible for 20% of all infections.

It is well recognized in statistical literature that the distribution underlying a data generating mechanism imposes a certain mean-to-variance relationship which in practice may be severely violated [18]. Despite this, the use of other distributions in infectious disease modelling that may just as well account for variation in disease transmission has been rather limited. Some studies suggest that SSEs follow a power-law distribution with fat tails, such as the generalized Pareto distribution [19]. Brooks-Pollock *et al*. [20] have used a negative binomial as well as a Poisson-lognormal distribution to model the distribution of cluster sizes for tuberculosis in the UK and the Netherlands. In this study, the Poisson-lognormal distribution provided a better fit to the UK data, indicating the importance of comparing different assumptions about the underlying distribution when variation in disease transmission is present.

To our knowledge, there are no studies that have explicitly investigated the possible bias in using the negative binomial distribution as an approximation to the underlying transmission process. We argue that it is important to compare different distributional assumptions since different distributions could portray different tail behaviour, and hence capture SSEs differently. In this work we explore the use of other Poisson mixture distributions for inference of the offspring mean and the amount of heterogeneity in disease transmission. We focus on the three-parameter generalized Gamma distribution for the individual reproduction number, because of its flexibility and the fact that it has as special cases the Gamma, Weibull, and lognormal distribution [21]. First, we carry out a simulation study to investigate the potential bias in the estimation of the offspring mean and its variance when the distribution that is fit to the data does not correspond to the actual data generating distribution. Next, we use the proposed distributions to (re-)analyze several COVID-19 datasets and investigate the impact of the considered offspring distribution on the estimation of the proportion of cases that is responsible for 80% of transmission, *p*_80%_.

## Results

### Simulation study

We consider the Poisson-generalized Gamma (POGG) and three submodels for the offspring distributions: negative binomial (NB), Poisson-lognormal (POLN), and Poisson-Weibull (POWB). See Methods for a description of these Poisson mixture distributions and how the simulation study was performed. In general we find that when overdispersion increases, estimates tend to become more biased when the considered offspring distribution does not correspond to the data generating distribution (Suppl. Table B.1). This is especially the case when considering estimates of the standard deviation. As overdispersion increases, the true distribution is more often considered as the best fit based on AIC. In particular, when the data generating mechanism deviates from the NB model, assuming the NB model will often lead to an underestimation of the standard deviation. Where results are missing, no estimates could be obtained.

### Expected versus realized proportions of transmission

Based on the estimated mean and variance of the considered offspring distribution, we can obtain estimates for the proportion of cases responsible for a certain amount of transmission. There are two different approaches for obtaining these proportions (see Methods), where one is based on the distribution of the individual reproduction number [3], and the other is based on the complete offspring distribution [14]. Here we show how the different offspring distributions can affect these proportions. Figure 1a shows the expected proportion of transmission due to the 20% most infectious cases for the varying levels of heterogeneity used in the simulation study, for the different offspring distributions. For all distributions, this proportion increases with an increasing amount of heterogeneity (i.e. higher *σ*, which for the negative binomial results in lower *k*). Thus, less heterogeneity leads to a smaller proportion of transmission being attributed to the 20% most infectious cases. In case of high overdispersion, there is a substantial difference in the expected proportions between the distributions. Since for the Poisson-generalized Gamma distribution it is not possible to specify the parameters from a given mean and variance, we only estimated these proportions at the specified settings used in the simulation study, hence these are represented as dots instead of lines. Figure 1b shows the expected proportion of transmission due to a given proportion of infectious cases for the different levels of overdispersion. It can again be seen that the difference between these estimates across the different distributions increases when overdispersion increases (i.e. higher *σ*) and likewise there is a substantial difference in terms of the expected proportions of transmission. When taking into account the additional variation coming from the Poisson process, the same increase in the proportion of transmission due to the 20% most infectious cases is seen, again with substantial differences between the distributions when overdispersion increases (Fig. 1c and 1d). These results also hold when *R* > 1 (Suppl. Fig. A.2). The vertical bars in Fig. 1c represent the uncertainty in the proportion of transmission due to the discrete nature of the offspring distribution. This should be interpreted as the range of transmission that will be due to the 20% most infectious cases. When taking into account this uncertainty surrounding these point estimates, a substantial difference is seen between the Poisson-lognormal and the other distributions for higher levels of overdispersion.

**Figure 1:**
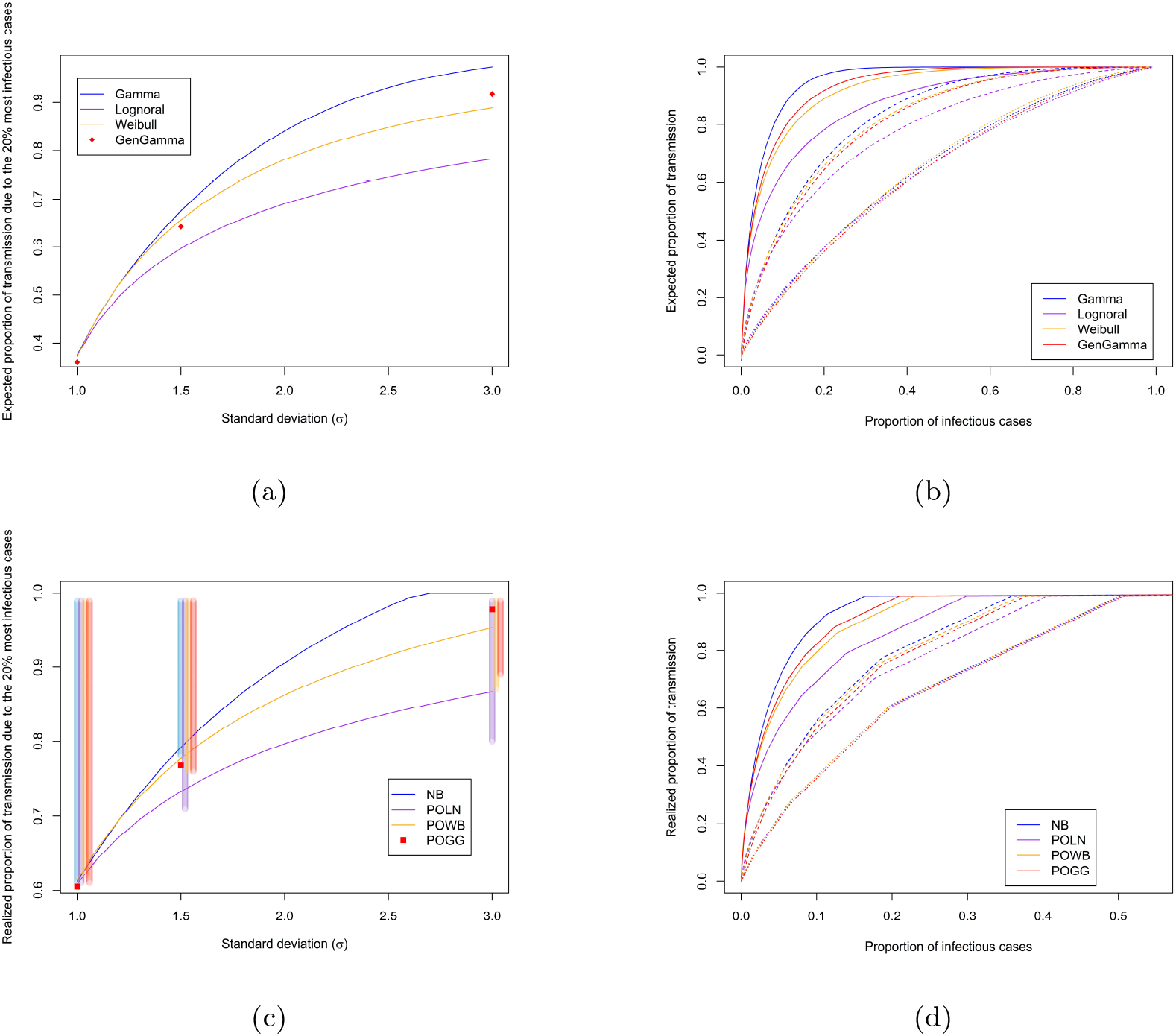
The top panel shows the expected proportion of all transmission that is (a) due to the 20% most infectious cases for different levels of overdispersion and different distributions, with the offspring mean *R* fixed at 0.8; and (b) due to a given proportion of infectious cases, where cases are ranked by their transmission potential, for *σ* = 1 (dotted), *σ* = 1.5 (dashed), *σ* = 3 (full), and the different distributions, with *R* fixed at 0.8. The lower panel shows the realized proportion of all transmission that is (c) due to the 20% most infectious cases, shaded vertical bars show the uncertainty surrounding the proportions at *σ* = {1, 1.5, 3}; and (d) due to a given proportion of infectious cases.

### Application to COVID-19 data

Table 1 shows estimates of the offspring mean *R* and standard deviation *σ* obtained by fitting the different offspring distributions to each COVID-19 dataset. In terms of AIC, in all cases the data are best described by a Poisson-lognormal distribution. Supplementary Figs. C.1, C.3, and C.5 show the fit of the different distributions to the observed offspring distribution. It can be seen that for the data from Hong Kong and India the negative binomial, Poisson-Weibull, and Poisson-generalized Gamma distributions underestimate the proportion of cases that generate only one secondary case, while this is captured well by the Poisson-lognormal distribution. For the data from Rwanda this is less evident. Goodness-of-fit plots are shown in Supplementary Figs. C.2, C.4, and C.6. For each of the three datasets (Table 1), *p*_80%_ is estimated to be substantially higher for the Poisson-lognormal than for the other distributions when based on the distribution of the individual reproduction number (Eq. (1), see Methods), and slightly higher when taking into account additional random variation from the Poisson process (Eq. (2), see Methods). Figure 2 shows these expected (left) and realized (right) proportions of transmission due to a certain proportion of cases for each distribution and each dataset. For Hong Kong, roughly 12-31% of cases are responsible for 68-100% of all transmission based on the Poisson-lognormal distribution (Fig. 2b). Based on the negative binomial distribution, roughly 14-31% of cases are responsible for 71-100% of all transmission.

**Table 1:** Estimates of the offspring mean *R* and its standard deviation (*σ*) using the different mixture distributions, and their AIC value, for three COVID-19 datasets. *p*_80%_ represents the proportion of cases responsible for 80% of transmission, following Equations (1) and (2). Estimates based on the negative binomial distribution correspond to those reported in the literature for the two published datasets.

| Dataset | Distribution | $R$ (95%CI) | $\sigma$ (95%CI) | AIC | $p_{80\%}$ | |
| --- | --- | --- | --- | --- | --- | --- |
|  |  |  |  |  | Eq. (1) | Eq. (2) |
| Hong Kong [13] | NB | 0.583 (0.448 - 0.718) | 1.175 (0.944 - 1.490) | 593.925 | 0.288 | 0.191 |
|  | POLN | 0.587 (0.456 - 0.779) | 1.413 (0.969 - 2.442) | 590.009 | 0.332 | 0.195 |
|  | POWB | 0.580 (0.445 - 0.745) | 1.218 (0.970 - 1.734) | 591.747 | 0.294 | 0.189 |
|  | POGG | 0.580 (0.3789 - 0.724) | 1.258 (0.923 - 1.550) | 592.738 | 0.303 | 0.190 |
| India [15] | NB | 0.484 (0.480 - 0.494) | 0.973 (0.962 - 0.985) | 163974.5 | 0.319 | 0.191 |
|  | POLN | 0.484 (0.477 - 0.491) | 1.077 (1.055 - 1.101) | 162980.6 | 0.373 | 0.195 |
|  | POWB | 0.483 (0.476 - 0.489) | 0.997 (0.984 - 1.011) | 163530.8 | 0.322 | 0.189 |
|  | POGG | 0.484 (0.477 - 0.490) | 1.012 (1.000 - 1.024) | 163286.5 | 0.333 | 0.191 |
| Rwanda | NB | 0.259 (0.216 - 0.302) | 0.623 (0.547 - 0.731) | 1015.261 | 0.323 | 0.138 |
|  | POLN | 0.260 (0.219 - 0.311) | 0.657 (0.560 - 0.820) | 1013.073 | 0.389 | 0.139 |
|  | POWB | 0.259 (0.217 - 0.311) | 0.631 (0.557 - 0.783) | 1014.350 | 0.331 | 0.137 |
|  | POGG | 0.259 (0.216 - 0.301) | 0.634 (0.561 - 0.706) | 1015.667 | 0.344 | 0.137 |

**Figure 2:**
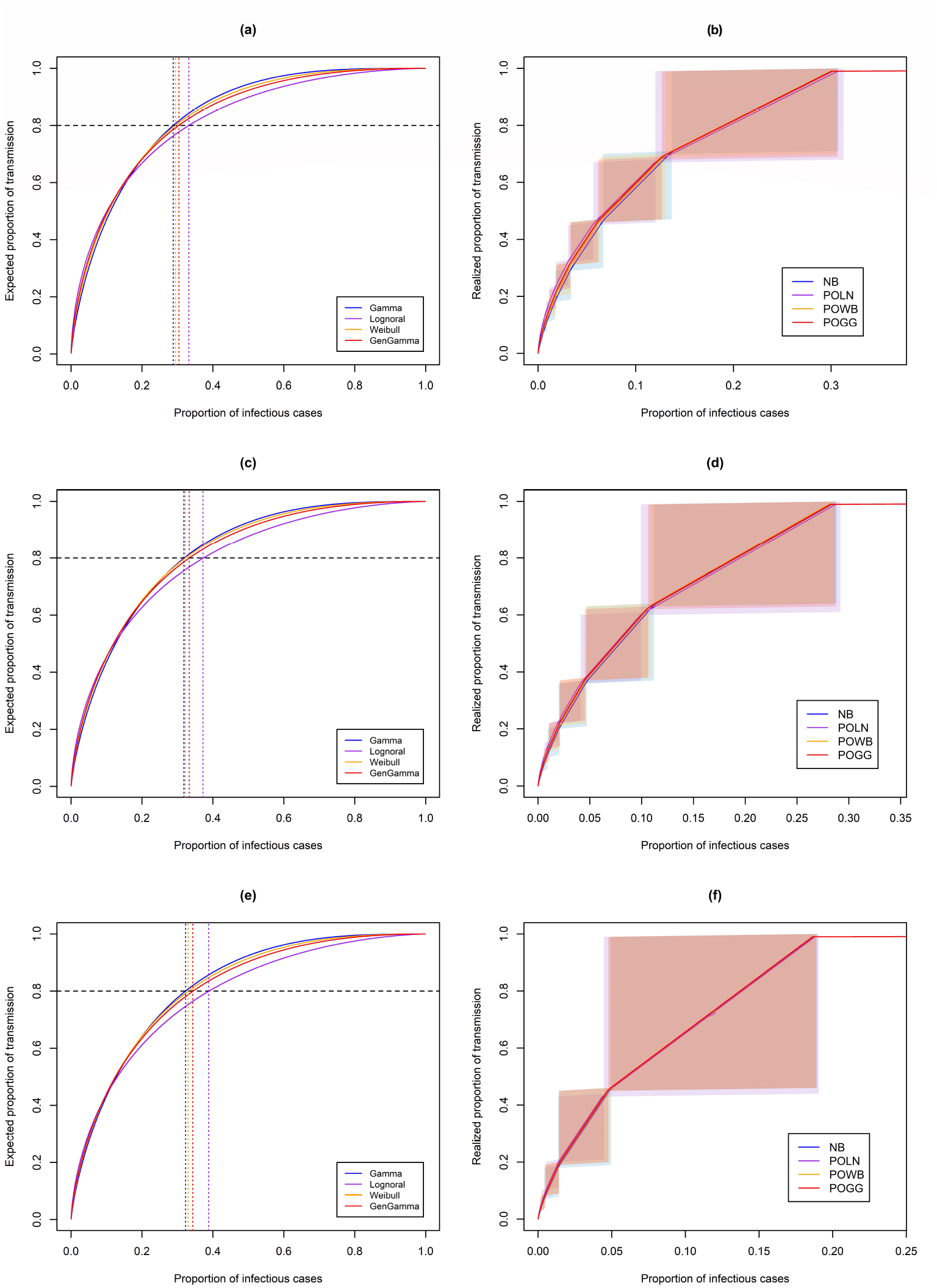
Proportion of most infectious cases responsible for a certain proportion of transmission, based on estimates from (a-b) Hong Kong, (c-d) India, and (e-f) Rwanda. Proportions are obtained based on the distribution of the individual reproduction number (left), and based on the complete offspring distribution (right). The shaded areas in the right panels represent the uncertainty surrounding specific proportions when considering the discrete nature of the realized offspring distributions.

For India, roughly 10-29% of cases are responsible for 61-100% of all transmission based on the Poisson-lognormal distribution (Fig. 2d). Based on the negative binomial distribution, roughly 11-29% of cases are responsible for 64-100% of all transmission. For Rwanda, roughly 5-19% of cases are responsible for 44-100% of all transmission based on the Poisson-lognormal distribution (Fig. 2f). Based on the negative binomial distribution, roughly 5-19% of cases are responsible for 46-100% of transmission.

## Discussion

Since most studies that aim to quantify variation in disease transmission have assumed the offspring distribution to follow a negative binomial, we investigated the impact of incorrectly assuming this distribution as an approximation to the underlying transmission process. Results from our simulation study show that when overdispersion increases, estimates of the offspring mean and especially its variance can become extremely biased when making incorrect assumptions about the underlying data generating distribution. When no variation in transmission is present, all distributions performed equally well, although there was a slightly increased bias in variance estimates when using the Poisson-Weibull or Poisson-generalized Gamma distribution. We have (re-)analyzed three COVID-19 datasets and in each case the Poisson-lognormal distribution gave the best fit to the observed data. This resulted in considerable differences in terms of the expected *p*_80%_ compared to when using a negative binomial distribution, when these proportions were based on the distribution of the individual reproduction number [3]. When accounting for the additional variation introduced by the Poisson process, the differences in these proportions of cases responsible became more subtle. For example, for the Hong Kong data the point estimate when using the Poisson-lognormal was 19.5%, compared to the previously reported 19.1% when using the negative binomial [13]. When accounting for the discrete nature of the offspring distribution, estimated ranges for these proportions were mostly overlapping for the different distributions, albeit a bit lower for the Poisson-lognormal distribution. Although most studies report *p*_80%_, the right-sided panels in Figure 2 indicate that depending on the proportion of transmission one is interested in, there might be a more substantial difference between the distributions. This implies that different distributions have different tail properties, underlining the importance of investigating which distribution best describes the data at hand. In addition we found that the difference between the distributions increases with an increasing level of overdispersion (Figure 1).

Our analyses indicated that the negative binomial distribution often underestimates the proportion of cases that generate only one secondary case, thereby possibly overestimating the importance of superspreading events. This overestimation was observed when the proportion of cases responsible was obtained based on the distribution for the underlying individual reproduction number. When accounting for the Poisson process, super-spreading was found to be only slightly more important when using a negative binomial distribution to describe the data, compared to a Poisson-lognormal distribution. A negative binomial distribution enables easy comparison between different studies through its dispersion parameter *k* [3]. However, this should not be a reason to only use negative binomial offspring distributions. The results from different studies can also be compared by their estimated *p*_80%_, which is often reported as well. It should be noted that there are different approaches for obtaining these proportions, hence care should be taken when comparing these results between studies. Lloyd-Smith *et al*. [3] assume SSEs to be realizations from the right-hand tail of the distribution of the individual reproduction number, hence their approach is based on this continuous distribution. In contrast, Endo *et al*. [14] have based these estimates on the complete offspring distribution, taking into account additional variation arising from the discrete Poisson process. In this way, the second approach accounts for more heterogeneity. However, if the effective contact process in reality is not a Poisson process, this approach may result in biased estimates.

In this study we have considered the three-parameter Poisson-generalized Gamma distribution, which has as special cases the Poisson-lognormal, Poisson-Weibull, and negative binomial distribution. Although the Poisson-generalized Gamma distribution has the advantage of being very flexible due to the additional parameter, the disadvantage is that because of this added complexity the estimation is computationally more extensive, especially for large datasets. Furthermore, parameter estimation can be difficult because different parameter sets can give rise to the same density function. In general, parameter estimation becomes more difficult when the amount of overdispersion is high and incorrect assumptions about the underlying data generating distribution are made. For that reason we were not able to fit the Poisson-Weibull and Poisson-lognormal distributions in some scenarios of our simulation study. This occurred when the data were highly overdispersed (*k* < 0.1), which is less likely to be encountered in practice [12].

Inference of the amount of heterogeneity in transmission is paramount for identifying a disease’s potential of superspreading. Correctly quantifying this heterogeneity is important because it affects estimates of other epidemiological parameters, modulates the degree of unpredictability of an epidemic, and needs to be taken into account when modeling disease control and planning control strategies [22]. When there is evidence of substantial superspreading, control measures should focus on limiting the potential for SSEs to occur by restricting large events and avoiding crowding in other public spaces. Typically, when control measures are taken, one aims to prevent transmission from those cases expected to have a high individual reproduction number, without knowing whether they will actually realize these secondary cases. Control measures thus act on the individual reproduction number and the expected transmissions, whereas their effect will be observed at the level of realized transmissions. Also, because of the increased speed at which the epidemic spreads when SSEs are present, heterogeneity could lower the doubling times [23].

Detailed contact tracing data are needed to obtain empirical offspring distributions, but these are often not available. Therefore this work should be extended such that the considered distributions can be used to infer the offspring mean *R* and its overdispersion from final size data [10], which are often more readily available. Rock *et al*. [24] have mentioned the distinction between ‘super-spreaders’ and ‘super-shedders’, who are both responsible for an above average number of secondary cases but for different reasons. In a meta-regression analysis, Chen *et al*. [25] investigated the relationship between the dispersion parameter *k* and respiratory viral load (rVL). They found that heterogeneity in rVL facilitates variation in individual infectiousness and hence may be associated with overdispersion in the number of secondary cases. Future work should thus aim to disentangle heterogeneity coming from variation in contact rates versus heterogeneity coming from variation in viral shedding. Furthermore, assuming a homogeneous Poisson process for effective contacts is likely a simplification of the real contact process, hence the use of other distributions to describe the contact process should also be considered. In a recent study, Wong & Collins [19] have suggested that the offspring distribution for SARS-CoV-2 is fat-tailed, which is consistent with a generalized Pareto distribution. We did not find evidence that a discrete Pareto offspring distribution would better describe the data used in this study (see Supplementary Text). Overall, the results of the present study suggest that, whenever possible, several distributions should be compared in terms of their fit to the observed data before making conclusions on the amount of heterogeneity by simply assuming a negative binomial offspring distribution.

## Methods

### Poisson mixture distributions

Differences in infectious disease transmission among individuals can arise either from differences in infectiousness or from differences in susceptibility, and can be interpreted in terms of the underlying contact and infection processes [26]. An effective contact is a contact that can lead to transmission, whereas an infectious contact occurs when an effective contact is realized between an infectious and susceptible individual. Effective contacts can be described using a Poisson counting process. Let *Y* denote the effective contact process that follows a Poisson distribution, *Y* ∼ *Po*(*ν*), where *ν* represents the individual reproduction number that itself is a random variable to allow for heterogeneity in transmission. The effective contact process *Y* is then described by a Poisson mixture distribution. In this work we focus on the three-parameter generalized Gamma distribution for *ν*, because of its flexibility and the fact that it has as special cases the Gamma, Weibull, and lognormal distribution [21]. Table 2 shows the resulting Poisson mixture distributions, each with mean *R* and variance *σ*^2^. More details can be found in the Supplementary Text.

**Table 2:** Different mixture distributions, assuming a Poisson distribution for the effective contact process.

| Distribution for $\nu$ | Offspring distribution | $R$ | $\sigma^2$ |
| --- | --- | --- | --- |
| $\nu \sim \text{Ga}(\alpha, \beta)$ | $Y \sim \text{NB}(\mu, k)$ | $\mu = \frac{\alpha}{\beta}$ | $\mu(1 + \frac{\mu}{k}) = \frac{\alpha}{\beta}(1 + \frac{1}{\beta})$ |
| $\nu \sim \text{LogN}(\mu_{log}, \sigma_{log})$ | $Y \sim \text{PoLN}(\mu_{log}, \sigma_{log})$ | $e^{\mu_{log} + \frac{\sigma_{log}^2}{2}}$ | $e^{\mu_{log} + \frac{\sigma_{log}^2}{2}} + [(e^{\sigma_{log}^2} - 1)e^{2\mu_{log} + \sigma_{log}^2}]$ |
| $\nu \sim \text{Weibull}(p, l)$ | $Y \sim \text{PoWB}(p, l)$ | $l\Gamma(1 + \frac{1}{p})$ | $l\Gamma(1 + \frac{1}{p}) + l^2[\Gamma(1 + \frac{2}{p}) - (\Gamma(1 + \frac{1}{p}))^2]$ |
| $\nu \sim \text{GG}(a, d, p)$ | $Y \sim \text{PoGG}(a, d, p)$ | $a \frac{\Gamma(\frac{d+1}{p})}{\Gamma(\frac{d}{p})}$ | $a \frac{\Gamma(\frac{d+1}{p})}{\Gamma(\frac{d}{p})} + a^2 \left[ \frac{\Gamma(\frac{d+2}{p})}{\Gamma(\frac{d}{p})} - \left( \frac{\Gamma(\frac{d+1}{p})}{\Gamma(\frac{d}{p})} \right)^2 \right]$ |

### Simulation study

To assess the possible bias in estimates of the reproduction number *R* and its overdispersion, which are based on assuming a certain offspring distribution for the underlying transmission process, we investigate the influence of the choice of distribution on the corresponding estimates. Using each of the mixtures in Table 2 we generate 1,000 datasets containing the distribution of secondary cases for 10,000 individuals. We set the mean number of secondary cases (i.e. the offspring mean) to 0.8, and vary the standard deviation *σ* = {1, 1.5, 3} corresponding to different levels of overdispersion (negative binomial *k* = {3.2, 0.44, 0.08}). We also consider a scenario without heterogeneity where the data are generated from a Poisson distribution with variance equal to the mean, *R* = *σ*^2^ = 0.8. We then estimate the parameters of the mixture distributions for each simulated dataset (*i* = 1, …, 1000) using maximum likelihood estimation (MLE) and obtain the estimated mean 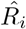 and standard deviation 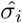 of the offspring distribution. For each distribution we calculate the bias in the estimates as 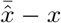, where *x* is the true value of the parameter of interest and 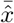 is the sample mean. Following Burton *et al*. [27], a bias larger than 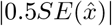 is alarming, where 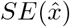 is the empirical standard error of the estimate 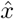 across all simulated datasets (i.e. the between-sample variability). We also obtain the bias as a percentage of the 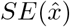, which ideally would be smaller than 40% in either direction [28]. Further, we calculate the mean squared error (MSE) as a measure of overall accuracy by taking into account the bias as well as the variability in the estimates. For example, a more flexible model such as the Poisson-generalized Gamma distribution is expected to have lower bias, but as a consequence of its complexity the variability is expected to be higher [29].

### Expected versus realized proportions of transmission

After estimating the mean 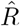 and variance 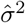 of the considered mixture distribution, we can obtain the proportion of cases responsible for a given proportion of transmission. Following Lloyd-Smith *et al*. [3], the parameters 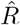 and 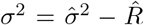 specify the probability density function (pdf) *f*_*ν*_ (*x*) and cumulative distribution function (cdf) *F*_*ν*_ (*x*) of the distribution describing the individual reproduction number *ν*. The cdf for disease transmission is defined by

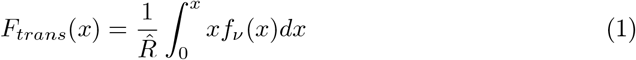

and denotes the expected proportion of transmission due to infectious cases with *ν* < *x*, while 1 − *F*_*trans*_(*x*) denotes the expected proportion of transmission due to those cases with *ν* > *x*. If *p* is the proportion of transmission for which we want to know the expected proportion of cases responsible, *a*, we first need to find *x* such that 1 − *F*_*trans*_(*x*) = *p*. The value *x* then denotes the threshold value of the reproduction number for which 1 − *F*_*trans*_(*x*) is the expected proportion of transmission *p* due to cases with *ν* > *x*. We can then obtain the expected proportion of cases which have their reproduction number *ν* > *x* as *P* (*X* > *x*) = 1 − *P* (*X* ≤ *x*) = 1 − *F*_*ν*_ (*x*). This is the expected proportion of infectious cases *a* that is responsible for a proportion *p* of all transmission. Note that in case of a homogeneous Poisson process the relation between *a* and *p* will be linear (Supplementary Fig. A.1b) because the variance of the mixing distribution will be zero.

If we want to take into account the additional variation coming from the Poisson process, we need to extend the method above for use with the Poisson mixtures (i.e. the offspring distributions). Endo *et al*. [14] have done this for the negative binomial distribution and we extend this for the other mixtures in the following way. The cdf for disease transmission is now defined by

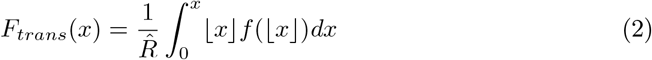

where *f* (|*x*|) is the density function of the mixture distribution evaluated at the integer part of *x. F*_*trans*_(*x*) now denotes the proportion of transmission that is due to cases that have their number of secondary cases *r* < *x*. Again we first need to find *x* such that 1 − *F*_*trans*_(*x*) = *p*, where *x* then denotes the threshold value of the reproduction number for which 1 − *F*_*trans*_(*x*) is the proportion of transmission *p* due to cases with *r* ≥ *x*. The proportion of cases that have *r* < *x* is defined as

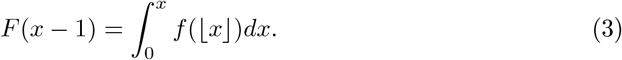

The proportion of cases that have their number of secondary cases *r* ≥ *x* is then *P* (*X* ≥ *x*) = *P* (*X* > *x*−1) = 1−*F* (*x*−1). This is now the proportion of cases *a* that is responsible for a proportion *p* of all transmission. However, as this is a continuous approximation of a discrete distribution, we should account for uncertainty in these point estimates of *a* and *p*. To do this, we use a discrete version of the method proposed by Lloyd-Smith *et al*. [3] (see details in Supplementary Text). We then obtain a range for the proportion of cases *a* responsible for a certain proportion of transmission *p*, which is then also expressed as a range.

More details on the difference between these two approaches can be found in Supplementary Text. Essentially, Lloyd-Smith *et al*. [3] estimate *the expected* proportion of cases responsible, whereas Endo *et al*. [14] estimate *the realized* proportion. We investigate the impact of the assumed offspring distribution on estimates of the proportion of infectious cases responsible for a certain amount of transmission.

### Application to COVID-19 data

Using MLE, we fit the different Poisson mixture distributions to three datasets containing the distribution of secondary cases for COVID-19. From the estimated parameters we calculate the mean *R* and standard deviation *σ* of the offspring distribution, and obtain their 95% confidence intervals (CI) by sampling 100,000 values from a multivariate normal distribution for the parameters of the offspring distribution. We compare the models in terms of AIC and goodness of fit based on observed vs. expected distribution of secondary cases. We also investigate the impact of the different distributions on the inference of *p*_80%_. We use two publicly available datasets, one containing the offspring distribution for 290 cases in Hong Kong [13], and one containing the offspring distribution for 84,965 cases in India [15]. The third dataset contains the offspring distribution for 795 cases in Rwanda (personal communication).

## Data Availability

The data from Hong Kong and India used in this study are publicly available. The empirical offspring distribution from Rwanda and the code to generate data as used in the simulation study, as well as all R code used in the analyses, are available on GitHub (https://github.com/cecilekremer/PoiMixtSS).

## Data availability

The data from Hong Kong and India used in this study are publicly available. The empirical offspring distribution from Rwanda and the code to generate data as used in the simulation study are available on GitHub (https://github.com/cecilekremer/PoiMixtSS).

## Code availability

All relevant R code used in this study is available on GitHub (https://github.com/cecilekremer/PoiMixtSS).

## Acknowledgements

The authors thank Muhammed Semakula for sharing the data obtained from Rwandan contact tracing efforts. This project has received funding from the European Union’s Horizon 2020 research and innovation programme - project EpiPose (Grant agreement number 101003688). The computational resources and services used in this work were provided by the VSC (Flemish Supercomputer Center), funded by the Research Foundation Flanders (FWO) and the Flemish Government department EWI. AT acknowledges support from the special research fund of the University of Antwerp. NH and AT acknowledge funding from the European Research Council (ERC) under the European Union’s Horizon 2020 research and innovation programme (grant agreement 682540 -TransMID).

## Author contributions

NH, CF, CK, and AT contributed to the conceptualization of the study. CK, AT, NH, SB, HM, and SV conducted the literature search. CK, AT, SB, HM, and SV contributed to the analysis code; CK and AT performed the analysis. NH, CF, CK, AT, and CA contributed to the interpretation of results. CK and AT drafted the manuscript. All co-authors critically reviewed and revised the manuscript. CK and AT contributed equally to this work.

## Competing interests

All authors have declared no competing interests.

## Notes

### Competing Interest Statement

The authors have declared no competing interest.

