## Supplementary Material for "Quantifying superspreading for COVID-19 using Poisson mixture distributions"

#### A Supplementary Text

##### Poisson mixture distributions

Let $Y$ denote the effective contact process, which is a combination of the Poisson contact process and the distribution of individual reproduction numbers. When assuming a constant rate $R$ for the transmission process, $Y$ follows a Poisson distribution with constant rate, $Y \sim Po(R)$. When accounting for individual variation, the rate itself is assumed to follow a distribution $(\nu)$, leading to a Poisson mixture distribution $Y \sim Po(\nu)$.

The variance of a mixture distribution $Y$ is given by $Var(Y) = E(Y^2) + \left[E(Y)\right]^2$. Using the probability generating function $Q(s)$ we can obtain the variance for each mixture, following

$$E(Y) = Q^{(1)}(1)$$
$$E(Y^2) = Q^{(1)}(1) + Q^{(2)}(1),$$

where $Q^{(n)}(1)$ is the $n^{th}$ derivative of $Q(s)$, evaluated at $s = 1$. If the individual reproduction numbers $\nu$ follow a distribution with pdf $f_\nu(\lambda)$, then the PGF of the Poisson mixture distribution is given by

$$Q(s) = \int_0^\infty e^{-\lambda(1-s)} f_\nu(\lambda) d\lambda. \tag{1}$$

Alternatively, for these Poisson mixture distributions, the variance equals the sum of the mean and variance of the mixing distribution $f_\nu(\lambda)$ [1]. Let $X$ be a random variable and define $Y$ as $Y|X = y \sim Po(x)$. Then using the law of total expectation,

$$E[Y] = E[E[Y|X]] = E[X]$$

since $E[Y|X] = X$, the variance can be obtained as

$$Var(Y) = E[Var(Y|X)] + Var(E[Y|X]) = E[X] + Var(X).$$

We consider different distributions for $\nu$, resulting in different mixture distributions for $Y$, each with mean $R$:

- $\nu$ is constant $(R) \rightarrow Y \sim$ Poisson$(R)$
- $\nu \sim$ Gamma with shape $\alpha$ and rate $\beta \rightarrow Y \sim$ Negative-Binomial$(\mu, k)$ with mean $\mu$ and dispersion parameter $k$
- $\nu \sim$ Weibull with shape $p$ and scale $l \rightarrow Y \sim$ Poisson-Weibull$(p, l)$ with shape $p$ and scale $l$
- $\nu \sim$ Lognormal with mean $\mu_{log}$ and standard deviation $\sigma_{log} \rightarrow Y \sim$ Poisson-lognormal$(\mu_{log}, \sigma_{log})$ with mean $\mu_{log}$ and standard deviation $\sigma_{log}$
- $\nu \sim$ generalized-Gamma with scale $a$ and shape parameters $d$ and $p \rightarrow Y \sim$ Poisson-generalized Gamma$(a, d, p)$ with scale $a$ and shape parameters $d$ and $p$

In the following sections we will describe the different mixture distributions $Y \sim$ Poisson$(\nu)$ that will be used (in this notation $x$ represents the number of secondary cases).

**Negative binomial**

The negative binomial distribution is a Poisson mixture where the individual reproduction number $\nu$ follows a Gamma distribution, i.e. $\nu \sim Ga(\alpha, \beta)$ with shape $\alpha$ and rate $\beta$. The density function of the negative binomial distribution is given by

$$f(x; \alpha, \beta) = \frac{\beta^\alpha}{\Gamma(\alpha) x!} \cdot \frac{\Gamma(x + \alpha)}{(1 + \beta)^{x+\alpha}} \tag{2}$$

where the mean is given by $\mu = \alpha/\beta$. The probability generating function of the negative binomial distribution [2] is given by

$$Q(s) = \left(1 + \frac{\mu}{k}(1 - s)\right)^{-k} \tag{3}$$

and hence the variance is $\mu(1 + \frac{\mu}{k}) = \frac{\alpha}{\beta}(1 + \frac{1}{\beta})$, and the dispersion parameter $k$ is given by $\frac{\mu^2}{\sigma^2 - \mu}$.

**Poisson-lognormal**

Consider a Poisson mixture where $\nu \sim$ Log-$N(\mu_{log}, \sigma_{log})$ where $\sigma_{log}$ and $\mu_{log}$ are the standard deviation and mean on the log-scale. The density function of the Poisson-lognormal distribution is given by

$$f(x; \mu_{log}, \sigma_{log}) = \int_0^\infty \frac{e^{-\lambda}\lambda^x}{x!} \cdot \frac{1}{\lambda \cdot \sigma_{log} \cdot \sqrt{2\pi}} e^{-\frac{(ln(\lambda)-\mu_{log})^2}{2\cdot\sigma_{log}^2}} d\lambda \quad (4)$$

where the mean is given by $e^{\mu_{log}+\frac{\sigma_{log}^2}{2}}$. The probability generating function is given by

$$Q(s) = \frac{e^{-\frac{\mu_{log}^2}{2\sigma_{log}^2}}}{\sigma_{log}\sqrt{2\pi}} \int_0^\infty \frac{1}{\lambda} e^{-\lambda(1-s)-\frac{log(\lambda)^2-2\mu_{log}log(\lambda)}{2\sigma_{log}^2}} d\lambda \quad (5)$$

and hence the variance is $e^{\mu_{log}+\frac{\sigma_{log}^2}{2}} + \left[(e^{\sigma_{log}^2}-1)e^{2\mu_{log}+\sigma_{log}^2}\right]$.

**Poisson-Weibull**

Consider a Poisson mixture where $\nu \sim$ Weibull$(p, l)$ with shape $p$ and scale $l$. The density function of the Poisson-Weibull distribution is given by

$$f(x; p, l) = \frac{p}{x! \cdot l^p} \int_0^\infty e^{-\lambda-(\frac{\lambda}{l})^p} \lambda^{x+p-1} d\lambda \quad (6)$$

where the mean is given by $l\Gamma(\frac{1}{p}+1)$. The probability generating function is given by

$$Q(s) = \int_0^\infty \frac{p}{l^p} e^{-\lambda(1-s)-(\frac{\lambda}{l})^p} \lambda^{p-1} d\lambda \quad (7)$$

and hence the variance is $l\Gamma(\frac{1}{p}+1) + l^2\left[\Gamma(1+\frac{2}{p}) - \left(\Gamma(1+\frac{1}{p})\right)^2\right]$.

**Poisson-generalized Gamma**

Consider a Poisson mixture where $\nu \sim$ GG$(a, d, p)$ where $a$ denotes the scale parameter and $d$ and $p$ denote the shape parameters. The density function of the Poisson-generalized Gamma distribution is given by

$$f(x; a, d, p) = \frac{p}{a^d \cdot x! \cdot \Gamma(\frac{d}{p})} \int_0^\infty \lambda^{x+d-1} \cdot e^{-(\frac{\lambda}{a})^p-\lambda} d\lambda \quad (8)$$

where the mean is given by $a\frac{\Gamma(\frac{d+1}{p})}{\Gamma(\frac{d}{p})}$. The probability generating function is given by

$$Q(s) = \int_0^\infty \frac{p}{a^d\Gamma(\frac{d}{p})} \lambda^{d-1} e^{-\lambda(1-s)-(\frac{\lambda}{a})^p} d\lambda \quad (9)$$

and hence the variance is $a\frac{\Gamma(\frac{d+1}{p})}{\Gamma(\frac{d}{p})} + a^2\left[\frac{\Gamma(\frac{d+2}{p})}{\Gamma(\frac{d}{p})} - \left(\frac{\Gamma(\frac{d+1}{p})}{\Gamma(\frac{d}{p})}\right)^2\right]$.

#### Expected vs. realized proportions of transmission

The approach used by Lloyd-Smith *et al.* [3] is based on the distribution of the individual reproduction number $\nu$, so it is only based on the inherent transmission potential (which is a combination of contacts and biological infectiousness) of individuals, not taking into account stochasticity in the transmission process. In this case, the expected proportion of cases responsible for a certain amount of transmission only depends on the level of overdispersion, and is more or less equal for each $R$. The approach used by Endo *et al.* [4] is based on the offspring distribution for the number of secondary cases, taking into account inherent individual variation as well as stochasticity in the transmission process. This can be interpreted as the *realized* proportion of cases responsible, as opposed to the *expected* proportion. In this case, the proportion responsible for a certain amount of transmission depends on the level of overdispersion as well as the reproduction number $R$ (Supplementary Fig. A.1a). Comparing the two approaches, Supplementary Fig. A.1b shows that the first approach results in lower expected proportions of transmission (i.e. a higher $p_{80\%}$), and the difference between the two approaches increases as the level of overdispersion decreases.

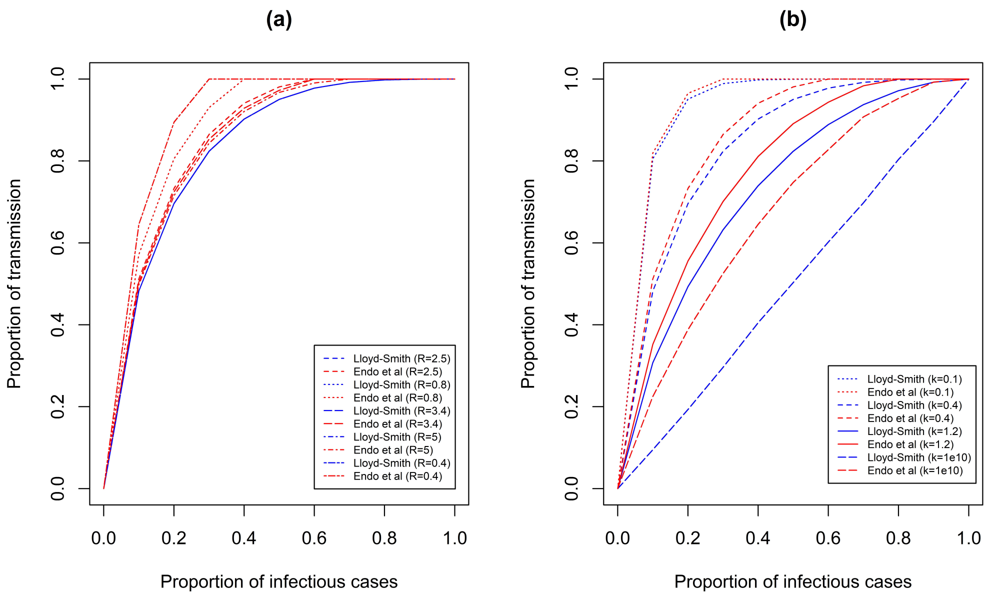


Figure A.1: Proportion of transmission due to a certain proportion of infectious cases for (a) varying $R$ with $k$ fixed at 0.4, and (b) varying levels of overdispersion $k$ and $R$ fixed at 2.5.

Essentially, the rate of the Poisson transmission process determines the overdispersion in the number of secondary cases $R$. This can either be a constant (no variation), or be taken from a distribution to allow for individual variation and hence create overdispersion. Suppose this rate is assumed to follow a Gamma distribution, then the result is a negative binomial offspring distribution with mean $R$ and dispersion parameter $k$. When the overdispersion level $k$ is very high ($\rightarrow \infty$), this indicates that there is no variation in the population (the variance of the Gamma distribution will be zero), which would imply the individual reproduction number to be constant. Then one would expect the relation between $a$ and $p$ to be linear when using Lloyd-Smith *et al.*'s approach [3]. When using the negative binomial distribution (i.e. the complete offspring distribution) to obtain the proportion responsible, this linear relation is not observed because it still accounts for variation due to the stochastic Poisson transmission process (Suppl. Fig. A.1b).

To account for the uncertainty in point estimates of $a$ and $p$ when these are based on the continuous approximation proposed by Endo *et al.* [4], we extend the method used by Lloyd-Smith *et al.* [3] by replacing the integral by a summation. Let $f(x)$ represent the probability mass function (pmf) of the offspring distribution and $F(x)$ the cdf of the offspring distribution. The cdf for disease transmission is then defined as

$$F_{trans}(x) = \frac{1}{R} \sum_{u=0}^{x} u f(u) \tag{10}$$

and denotes the proportion of transmission that is due to infectious cases with their number of secondary cases $r \leq x$. Again, we should find $x$ such that $1 - F_{trans}(x) = p$ where $x$ then denotes the threshold value of the reproduction number for which $1 - F_{trans}(x)$ is the proportion of transmission due to cases with $r > x$. The proportion of cases responsible for a proportion $p$ of transmission is then found as $P(X > x) = 1 - P(X \leq x) = 1 - F(x)$. However, in the discrete case it is unlikely that there exists an integer $x$ such that $1 - F_{trans}(x)$ exactly equals $p$. Therefore we need to define two values for this threshold, $x_1$ and $x_2$, such that $F_{trans}(x_1) < 1 - p < F_{trans}(x_2)$. The proportion of cases $a$ responsible for a range $p$ of transmission is then given by the range $\big[1 - F(x_2), 1 - F(x_1)\big]$.

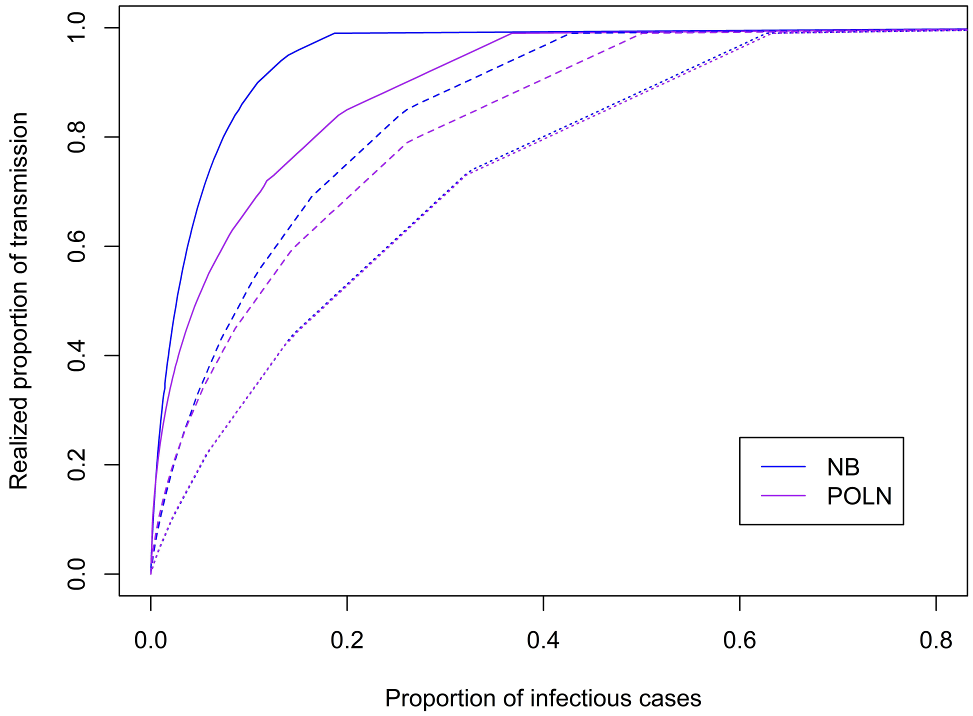


Figure A.2: Proportion of transmission due to a certain proportion of infectious cases for varying levels of overdispersion ($k = 0.08$ (full), $k = 0.44$ (dashed), $k = 3.2$ (dotted)) and $R$ fixed at 1.2, for the negative binomial vs. Poisson-lognormal distribution.

#### Discrete Pareto distribution

Wong & Collins [5] examined the tail behavior of the empirical offspring distribution $Z$, focusing on SSEs resulting in more than 6 secondary cases and found that this was inconsistent with exponential decay but rather exhibited fat-tail behavior. Based on these results they argue that the tail of $Z$ can be described by a generalized Pareto distribution.

For completeness, we investigate whether a discrete Pareto distribution [6] would perform better than the Poisson mixtures in describing the offspring distribution in our three data examples. The discrete Pareto distribution has probability mass function

$$f(x) = \theta^{log(1+x)} - \theta^{log(2+x)}$$

where $0 < \theta < 1$ is the shape parameter. We fit this distribution to each COVID-19 dataset and found that it does not provide a better fit in terms of AIC (Suppl. Table A.1) compared to the Poisson mixtures. It does seem to provide a slightly better fit in the tail for the India and Rwanda data, but does not adequately capture the proportion of cases that generate zero or only one secondary case (Suppl. Fig. C.7).

Table A.1: AIC values for the fit of a discrete Pareto distribution to each COVID-19 dataset.

| Dataset | AIC |
|---|---|
| Hong Kong | 597.141 |
| India | 166616.4 |
| Rwanda | 1030.718 |

### B Supplementary Table

Table B.1: Simulation study results. $R$ and $\sigma$ denote the offspring mean and standard deviation, respectively. $SE(\hat{x})$ is the between-sample variability of $\hat{x}$ and is used to obtain the standardized bias (Std. bias). Smaller MSE indicates higher accuracy. Best fit denotes the proportion of simulations where the fitted model has the lowest AIC. Asterisks indicate whether the bias is larger than $|0.5SE(\hat{x})|$.

| Data generating model | $\sigma$ ($k$) | Fitted model | Bias | | $SE(\hat{\beta})$ | | Std. bias | | MSE | | Best fit |
|---|---|---|---|---|---|---|---|---|---|---|---|
| | | | $\hat{R}$ | $\hat{\sigma}$ | $\hat{R}$ | $\hat{\sigma}$ | $\hat{R}$ | $\hat{\sigma}$ | $\hat{R}$ | $\hat{\sigma}$ | |
| Poisson | | NB | -0.0001 | 0.0025 | 0.0089 | 0.0062 | -0.8345 | 39.6070 | 0.0001 | 0.0001 | 0.3060 |
| | | PoLN | -0.0003 | 0.0025 | 0.0090 | 0.0064 | -3.2752 | 38.3755 | 0.00001 | 0.0001 | 0.1670 |
| | | PoWB | 0.0003 | 0.0031$^*$ | 0.0090 | 0.0061 | 3.4151 | 51.5682 | 0.0001 | 0.0001 | 0.4940 |
| | | PoGG | 0.0001 | 0.0032$^*$ | 0.0090 | 0.0063 | 1.1868 | 49.9982 | 0.0001 | 0.0001 | 0.0330 |
| NB | 1 (3.2) | NB | -0.0003 | -0.0001 | 0.0102 | 0.0110 | -3.1029 | -1.1811 | 0.0001 | 0.0001 | 0.5260 |
| | | PoLN | -0.0002 | 0.0015 | 0.0102 | 0.0114 | -2.1566 | 13.1819 | 0.0001 | 0.0001 | 0.1710 |
| | | PoWB | -0.0003 | -0.0020 | 0.0102 | 0.0109 | -3.0582 | -18.6363 | 0.0001 | 0.0001 | 0.2960 |
| | | PoGG | -0.0003 | -0.0008 | 0.0102 | 0.0115 | -3.2947 | -6.8577 | 0.0001 | 0.0001 | 0.0070 |
| | 1.5 (0.44) | NB | -0.0011 | -0.0024 | 0.0147 | 0.0268 | -7.6078 | -8.9771 | 0.0002 | 0.0007 | 0.8450 |
| | | PoLN | 0.0392$^*$ | 0.5053$^*$ | 0.0166 | 0.0671 | 235.8058 | 753.6438 | 0.0018 | 0.2598 | 0.0000 |
| | | PoWB | 0.0008 | 0.0462$^*$ | 0.0148 | 0.0308 | 5.1606 | 149.7747 | 0.0002 | 0.0031 | 0.0070 |
| | | PoGG | -0.0006 | -0.0036 | 0.0152 | 0.0305 | -3.8462 | -11.8622 | 0.0002 | 0.0009 | 0.1480 |
| | 3 (0.08) | NB | 0.0009 | 0.0042 | 0.0303 | 0.1158 | 2.8391 | 3.5873 | 0.0009 | 0.0134 | - |
| | | PoLN | 1.2265$^*$ | 130.5016$^*$ | 0.1873 | 26.5420 | 654.7167 | 491.6793 | 1.5393 | 17735.15 | - |
| | | PoWB | - | - | - | - | - | - | - | - | - |
| | | PoGG | 0.0745$^*$ | 0.1850$^*$ | 0.1252 | 0.2509 | 59.5224 | 73.7035 | 0.0212 | 0.0972 | - |
| PoLN | 1 (3.2) | NB | -0.0002 | -0.0037 | 0.0100 | 0.0109 | -1.6911 | -34.3200 | 0.0001 | 0.0001 | 0.2140 |
| | | PoLN | -0.0002 | -0.0006 | 0.0100 | 0.0114 | -1.5593 | -5.1894 | 0.0001 | 0.0001 | 0.7620 |
| | | PoWB | -0.0002 | -0.0056$^*$ | 0.0100 | 0.0109 | -1.4735 | -51.0669 | 0.0001 | 0.0002 | 0.0230 |
| | | PoGG | -0.0004 | -0.0024 | 0.0100 | 0.0112 | -3.6593 | -21.3130 | 0.0001 | 0.0001 | 0.0010 |
| | 1.5 (0.44) | NB | -0.0005 | -0.1585$^*$ | 0.0144 | 0.0283 | -3.7409 | -559.3826 | 0.0002 | 0.0259 | 0.0000 |
| | | PoLN | -0.0004 | -0.0002 | 0.0143 | 0.0497 | -2.8452 | -0.3733 | 0.0002 | 0.0022 | 0.9780 |
| | | PoWB | -0.0028 | -0.1252$^*$ | 0.0145 | 0.0323 | -18.9971 | -387.6966 | 0.0002 | 0.0167 | 0.0000 |
| | | PoGG | -0.0063 | -0.0912$^*$ | 0.0143 | 0.0365 | -43.9418 | -249.7971 | 0.0002 | 0.0097 | 0.0220 |
| | 3 (0.08) | NB | -0.0007 | -1.1417$^*$ | 0.0296 | 0.0860 | -2.4218 | -1327.6740 | 0.0009 | 1.3108 | 0.0000 |
| | | PoLN | -0.0017 | -0.0100 | 0.0262 | 0.2117 | -6.4958 | -4.7241 | 0.0007 | 0.0449 | 1.0000 |
| | | PoWB | -0.0339$^*$ | -1.0079$^*$ | 0.0201 | 0.0703 | -169.0068 | -1433.1900 | 0.0016 | 1.0209 | 0.0000 |
| | | PoGG | 0.0550 | -0.7524 | 0.3930 | 1.2907 | 13.9920 | -58.2917 | 0.1575 | 2.2321 | 0.0000 |
| PoWB | 1 (3.2) | NB | -0.0002 | -0.0010 | 0.0099 | 0.0107 | -1.7634 | -9.1319 | 0.0001 | 0.0001 | 0.2770 |
| | | PoLN | -0.00003 | -0.0005 | 0.0099 | 0.0110 | -0.3245 | -4.1545 | 0.0001 | 0.0001 | 0.0260 |
| | | PoWB | -0.0002 | -0.0029 | 0.0099 | 0.0105 | -1.7105 | -27.0875 | 0.0001 | 0.0001 | 0.5760 |
| | | PoGG | -0.0001 | -0.0025 | 0.0099 | 0.0106 | -0.7852 | -23.7951 | 0.0001 | 0.0001 | 0.1210 |
| | 1.5 (0.44) | NB | -0.0002 | -0.0423$^*$ | 0.0156 | 0.0284 | -1.5173 | -148.7519 | 0.0002 | 0.0026 | 0.0240 |
| | | PoLN | 0.0262$^*$ | 0.3438$^*$ | 0.0171 | 0.0615 | 153.8501 | 559.0264 | 0.0010 | 0.1220 | 0.0000 |
| | | PoWB | -0.0004 | -0.0009 | 0.0156 | 0.0319 | -2.3372 | -2.7970 | 0.0002 | 0.0010 | 0.8500 |
| | | PoGG | -0.0007 | -0.0026 | 0.0156 | 0.0340 | -4.3863 | -7.6866 | 0.0002 | 0.0012 | 0.1260 |
| | 3 (0.08) | NB | -0.0001 | -0.6792$^*$ | 0.0294 | 0.0976 | -0.3416 | -696.1316 | 0.0009 | 0.4708 | - |
| | | PoLN | - | - | - | - | - | - | - | - | - |
| | | PoWB | -0.0021 | -0.0194 | 0.0358 | 0.1794 | -5.9692 | -10.8080 | 0.0013 | 0.0325 | - |
| | | PoGG | 0.0247 | -0.2845 | 0.1120 | 0.3334 | 22.0196 | -85.3332 | 0.0132 | 0.1921 | - |
| PoGG | 1 (3.2) | NB | 0.0003 | 0.0004 | 0.0097 | 0.0106 | 2.8584 | 3.3602 | 0.0001 | 0.0001 | 0.4290 |
| | | PoLN | 0.0003 | 0.0013 | 0.0097 | 0.0109 | 3.5454 | 11.9935 | 0.0001 | 0.0001 | 0.1940 |
| | | PoWB | 0.0003 | -0.0013 | 0.0096 | 0.0105 | 2.8438 | -12.6182 | 0.0001 | 0.0001 | 0.3450 |
| | | PoGG | 0.0003 | -0.0001 | 0.0097 | 0.0105 | 2.9638 | -1.0649 | 0.0001 | 0.0001 | 0.0320 |
| | 1.5 (0.44) | NB | 0.0002 | -0.0365$^*$ | 0.0150 | 0.0279 | 1.1275 | -130.9008 | 0.0002 | 0.0021 | 0.0540 |
| | | PoLN | 0.0265$^*$ | 0.3350$^*$ | 0.0164 | 0.0605 | 161.2018 | 553.5758 | 0.0010 | 0.1159 | 0.0000 |
| | | PoWB | 0.0002 | 0.0020 | 0.0150 | 0.0314 | 1.3855 | 6.3352 | 0.0002 | 0.0010 | 0.8460 |
| | | PoGG | 0.0013 | -0.0050 | 0.0197 | 0.0357 | 7.7309 | -14.0632 | 0.0003 | 0.0013 | 0.1000 |
| | 3 (0.08) | NB | 0.0005 | -0.5106$^*$ | 0.0306 | 0.1068 | 1.6531 | -478.0705 | 0.0010 | 0.2721 | - |
| | | PoLN | - | - | - | - | - | - | - | - | - |
| | | PoWB | - | - | - | - | - | - | - | - | - |
| | | PoGG | -0.0006 | 0.0100 | 0.0835 | 0.4455 | -0.7113 | 2.2512 | 0.0070 | 0.1986 | - |

### C Supplementary Figures

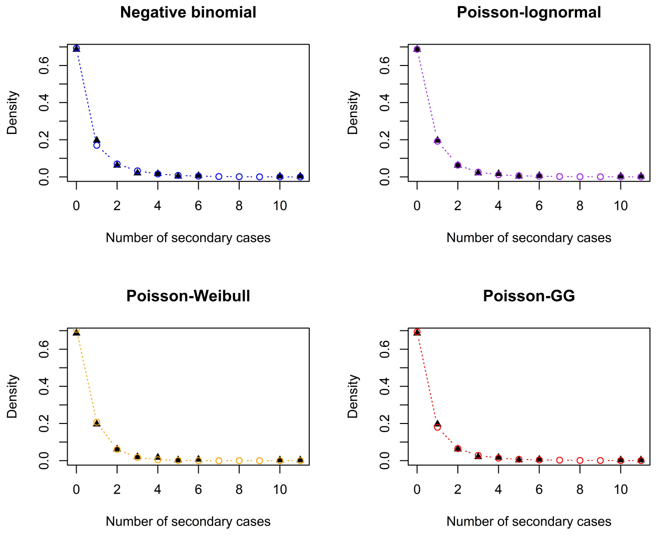


Figure C.1: Fit of the different distributions, **Hong Kong data** [7]. Triangles show the observed offspring distribution.

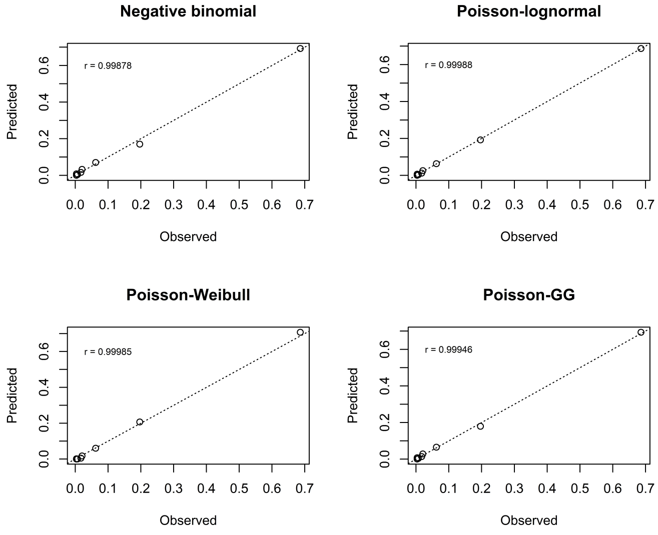


Figure C.2: Goodness-of-fit for the different distributions, **Hong Kong data** [7]. Deviations from the straight line indicate a lack of fit, with downward deviations indicating underestimation and upward deviations indicating overestimation. $r$ denotes the Pearson correlation coefficient.

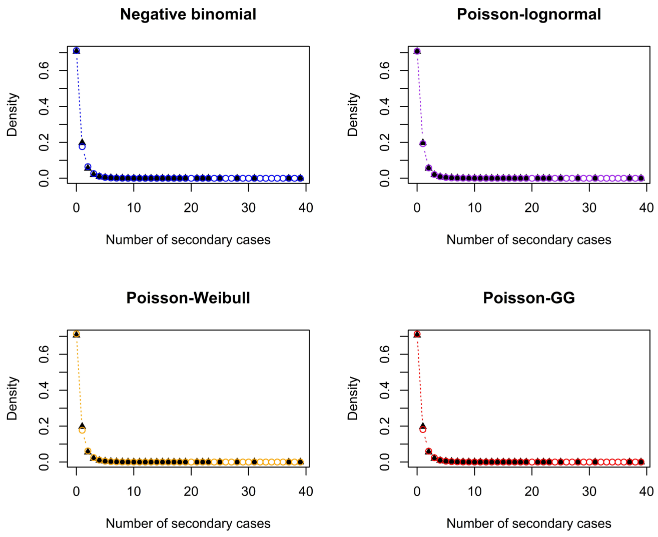


Figure C.3: Fit of the different distributions, **India data** [8]. Triangles show the observed offspring distribution.

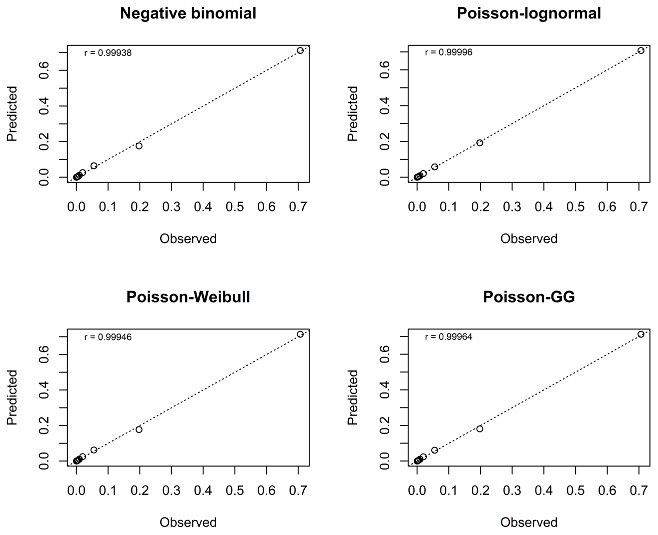


Figure C.4: Goodness-of-fit for the different distributions, **India data** [8]. Deviations from the straight line indicate a lack of fit, with downward deviations indicating underestimation and upward deviations indicating overestimation. $r$ denotes the Pearson correlation coefficient.

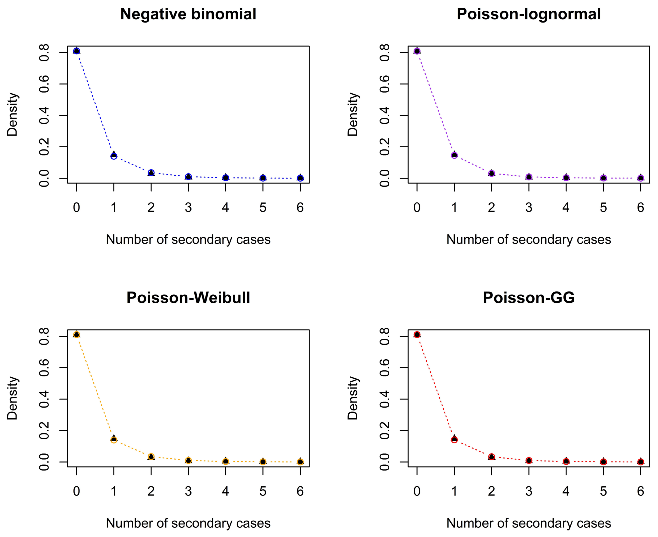


Figure C.5: Fit of the different distributions, **Rwanda data**. Triangles show the observed offspring distribution.

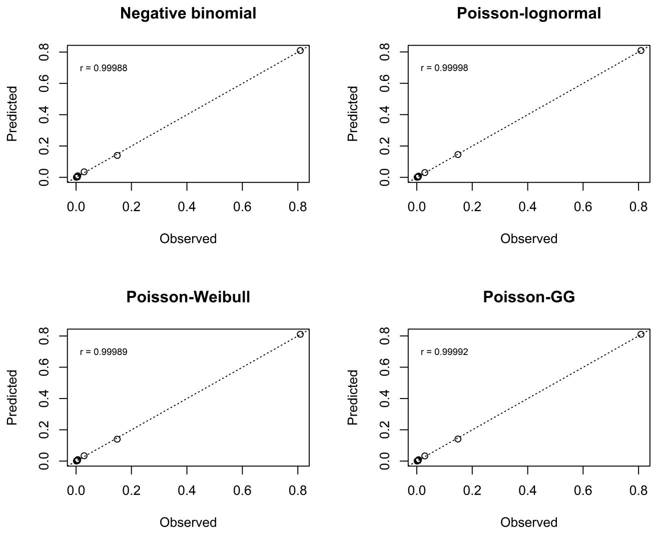


Figure C.6: Goodness-of-fit for the different distributions, **Rwanda data**. Deviations from the straight line indicate a lack of fit, with downward deviations indicating underestimation and upward deviations indicating overestimation. $r$ denotes the Pearson correlation coefficient.

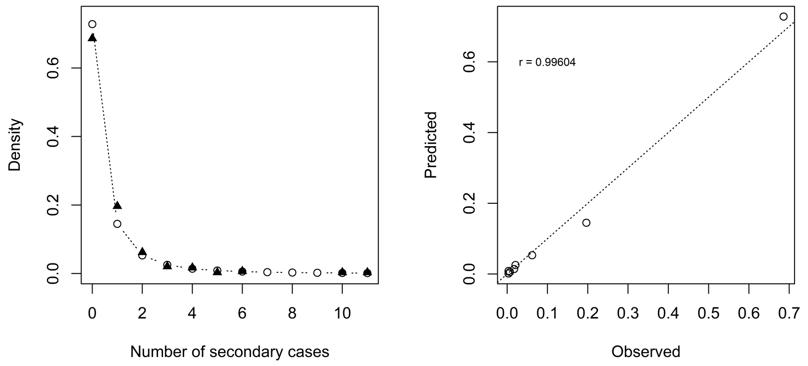


(a)

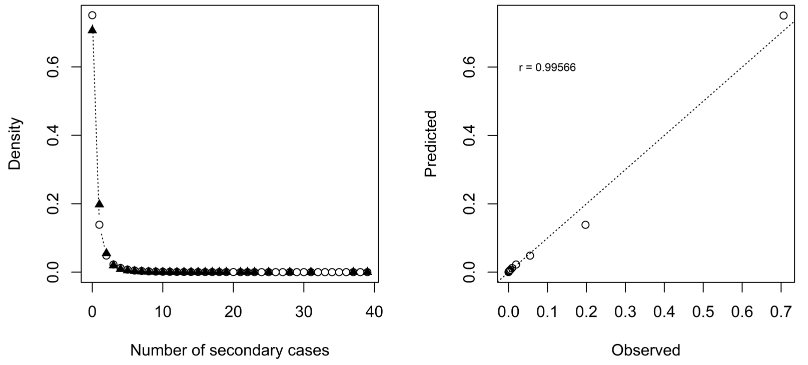


(b)

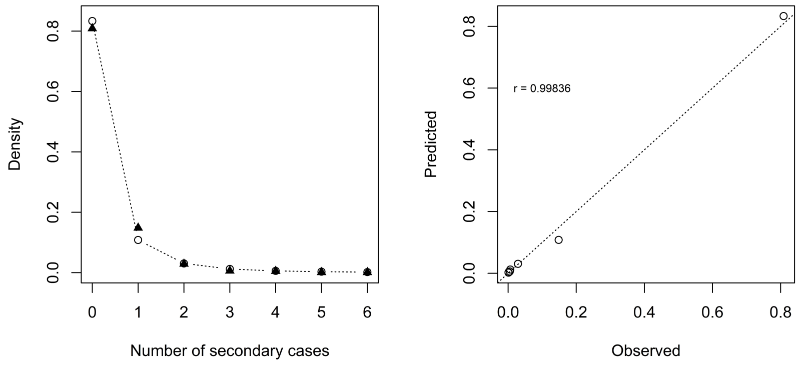


(c)

Figure C.7: Left panels show the fit of the discrete Pareto distribution to the observed (triangles) offspring distribution, right panels show the goodness-of-fit (deviations from the straight line indicate a lack of fit, $r$ denotes the Pearson correlation coefficient), for the data from (a) Hong Kong, (b) India, and (c) Rwanda.
